# RiskAgent: Autonomous Medical AI Copilot for Generalist Risk Prediction

**DOI:** 10.1101/2025.04.03.25323489

**Authors:** Fenglin Liu, Jinge Wu, Hongjian Zhou, Xiao Gu, Soheila Molaei, Anshul Thakur, Lei Clifton, Honghan Wu, David A. Clifton

## Abstract

The application of Large Language Models (LLMs) to various clinical applications has attracted growing research attention. LLMs currently achieve competitive results compared to human experts in examinations. However, real-world clinical decisionmaking differs significantly from the standardized, exam-style scenarios commonly used in current efforts. It therefore remains a challenge to apply LLMs to complex medical tasks that require a deep understanding of medical knowledge. A common approach is to fine-tune LLMs for target tasks, which, however, not only requires substantial data and computational resources but is also still prone to generating ‘hallucinations’. In this paper, we present the RiskAgent system to perform a broad range of medical risk predictions, covering over 387 risk scenarios across diverse complex diseases, e.g., cardiovascular disease and cancer. RiskAgent is designed to collaborate with hundreds of clinical decision tools, i.e., risk calculators and scoring systems that are supported by evidence-based medicine. To evaluate our method, we have built the first benchmark MedRisk specialized for risk prediction, including 12,352 questions spanning 154 diseases, 86 symptoms, 50 specialties, and 24 organ systems. The results show that our RiskAgent, with 8 billion model parameters, achieves 76.33% accuracy, outperforming the most recent commercial LLMs, o1, o3-mini, and GPT-4.5, and doubling the 38.39% accuracy of GPT-4o. On rare diseases, e.g., Idiopathic Pulmonary Fibrosis (IPF), RiskAgent outperforms o1 and GPT-4.5 by 27.27% and 45.46% accuracy, respectively. Finally, we further conduct a generalization evaluation on an external evidence-based diagnosis benchmark and show that our RiskAgent achieves the best results. These encouraging results demonstrate the great potential of our solution for diverse diagnosis domains. For example, instead of extensively fine-tuning LLMs for different medical tasks, our method, which collaborates with and utilizes existing evidence-based medical tools, not only achieves trustworthy results but also reduces resource costs, thus making LLMs accessible to resource-limited clinical applications. To improve the adaptability of our model in different scenarios, we have built and open-sourced a family of models ranging from 1 billion to 70 billion parameters. Our code, data, and models are all available at https://github.com/AI-in-Health/RiskAgent.

## Introduction

Large Language Models (LLMs), such as the GPT series ^1,2,3,4^ and PaLM ^5^, have shown promising performance in understanding text and assisting humans. Inspired by the success of LLMs, an increasing amount of research is attempting to apply LLMs to the medical field and explore their potential in assisting healthcare professionals, resulting in different types of medical LLMs ^6^. Specifically, based on the LLMs PaLM ^5^ and Gemini ^7^, Google has developed MedPaLM-2^8^ and Med-Gemini ^9,10^, respectively, which achieve 86.5% and 91.1% accuracy comparable to the 87.0% accuracy of human experts ^11^ on the US Medical Licensing Examination (USMLE). Based on open-source LLM LLaMA ^12,13^, dozens of medical LLMs have been proposed, such as Clinical Camel ^14^ and Meditron ^15^, to address different medical tasks.

However, the effective application of LLMs to complex medical tasks remains challenging: 1) **Clinical inefficacy**: LLMs achieve superior performance mainly in examination tasks ^17^. However, existing LLMs, including GPT-4, perform poorly on tasks closer to clinical decision-making, such as medical code querying ^18^ and new drug understanding ^17^. 2) **Resource-intensive**: To enable LLMs to deliver accurate results on different tasks, a potential solution is fine-tuning LLMs on target medical data ^19,6^. However, fine-tuning LLMs (forcing them to learn all tasks) not only places significant learning pressure on them, resulting in decreased performance, but also inevitably requires substantial data (ranging from 1 million ^20,21,8^ to 80 billion tokens ^15,11^) and computational resources that may be prohibitively difficult to obtain. More importantly, these fine-tuned medical LLMs do not utilize or collaborate with various tools already deployed in hospitals, including devices, models, and APIs, leading to resource waste. 3) **Privacy concern**: An alternative solution is to prompt commercial LLMs (e.g., the GPT series ^22^). However, the use of commercial LLMs involves strict privacy regulations regarding sensitive patient information in the real world, raising privacy concerns for their adoption in hospital scenarios ^23^. 4) **Unfaithful outputs**: LLMs still face the widely known ‘hallucination’ challenge ^24,25,26^: the produced answers appear reasonable but are not based on factual information and knowledge. Furthermore, existing LLMs could not effectively and accurately provide evidence to show the sources of generated answers, which is crucial in healthcare ^27,6^. Therefore, developing different medical LLMs for diverse medical tasks is expensive, time-consuming, and energy-intensive.

In this work, we present RiskAgent for solving complex medical problems and take risk prediction as a representative example that requires LLMs to not only accurately understand complex patients’ health records, but also predict potential health risks, including the risk of developing diseases or the mortality/survival rate of diseases. This is critical for preventative health ^28,29^, since early and accurate risk prediction can alert physicians and patients for early intervention, and thus improve clinical outcomes (e.g., increasing patient survival), especially for complex diseases such as cardiovascular disease and rare diseases ^30,31^. As shown in Figure 1(a), our RiskAgent includes three LLM agents: Decider, Executor, and Reviewer: 1) The Decider analyzes medical problems and accurately selects appropriate tools from hundreds of options; 2) The Executor understands the selected tools, parses their required parameters, and executes them; 3) The Decider then analyzes the returned tool outputs and provides initial answers; 4) The Executor structures and executes outputs to generate the final answers; 5) The Reviewer finally reviews the decision-making process and provides reflection on the results. To comprehensively evaluate LLMs’ risk prediction performance for diverse scenarios, we build a risk prediction benchmark MedRisk including 154 diseases, 86 symptoms, 50 specialties, and 24 organ systems, totaling 12,352 cases. Figure 1(b) presents the benchmark’s statistics. We evaluate 13 state-of-the-art methods, covering both general and medical LLMs, as well as open-source public and commercial LLMs. Figure 1(c) and Table 1 reveal that current LLMs perform poorly in making accurate risk predictions (15.83% ~58.77% accuracy) for complex diseases, while our RiskAgent substantially outperforms existing LLMs with large margins. Figure 1(d) shows that our method makes more accurate risk predictions than GPT-4o ^3^ for different complex diseases. The first two GPT-4o examples show that incorrectly predicting high-risk patients as low-risk would lead to delayed treatment. Conversely, when low-risk patients are predicted to be highrisk, as shown in the third GPT-4o example, it may cause over-treatment, waste medical resources, and bring unnecessary anxiety to patients.

**Table 1.**
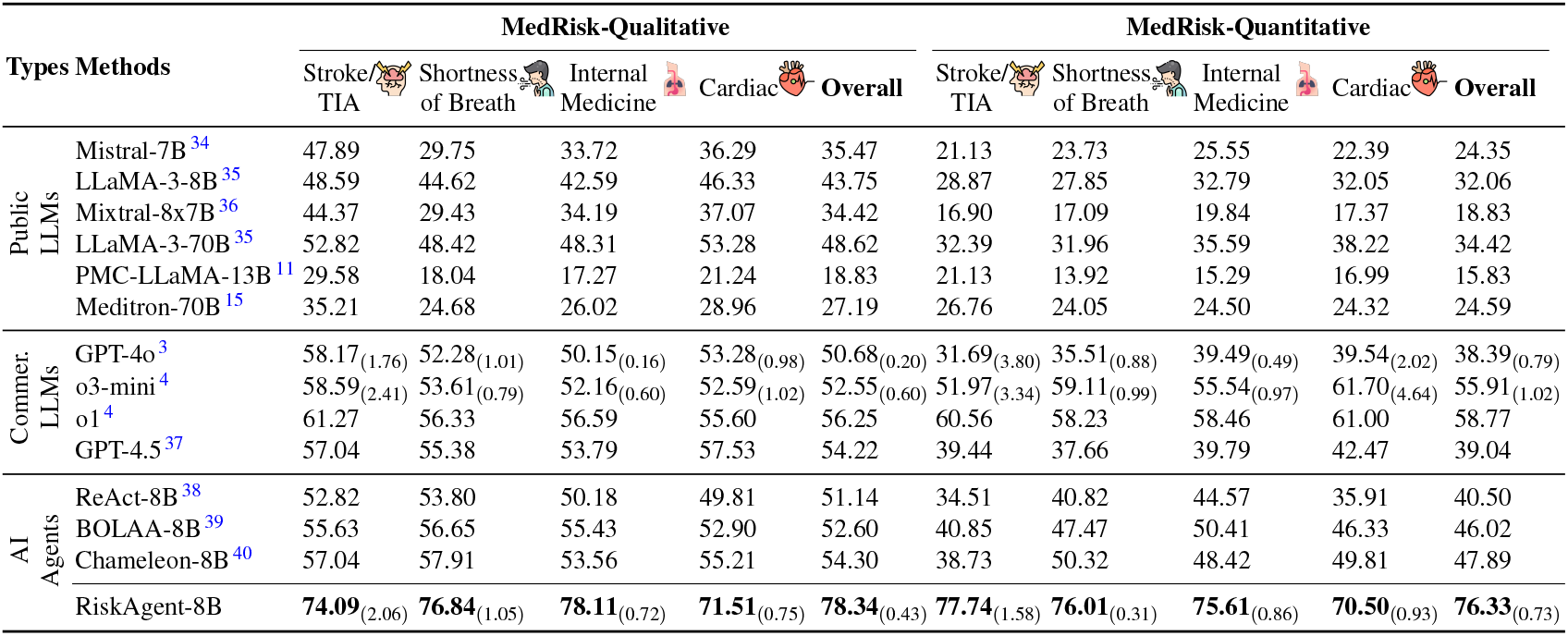
Performances of different methods on risk predictions. ‘B’: Billion. We use accuracy to report our results. We conduct five runs for our RiskAgent, GPT-4o, and o3-mini. We report the mean and standard deviation_(STD)_ of performance. In this paper, all values are reported in percentage (%). Higher is better for all columns. The bold number denotes the best result across all methods. We select the most common cases across disease, symptom, specialty, and organ system for a broad evaluation and subsequently report the overall results (i.e., the average performance on all test data).

**Figure 1.**
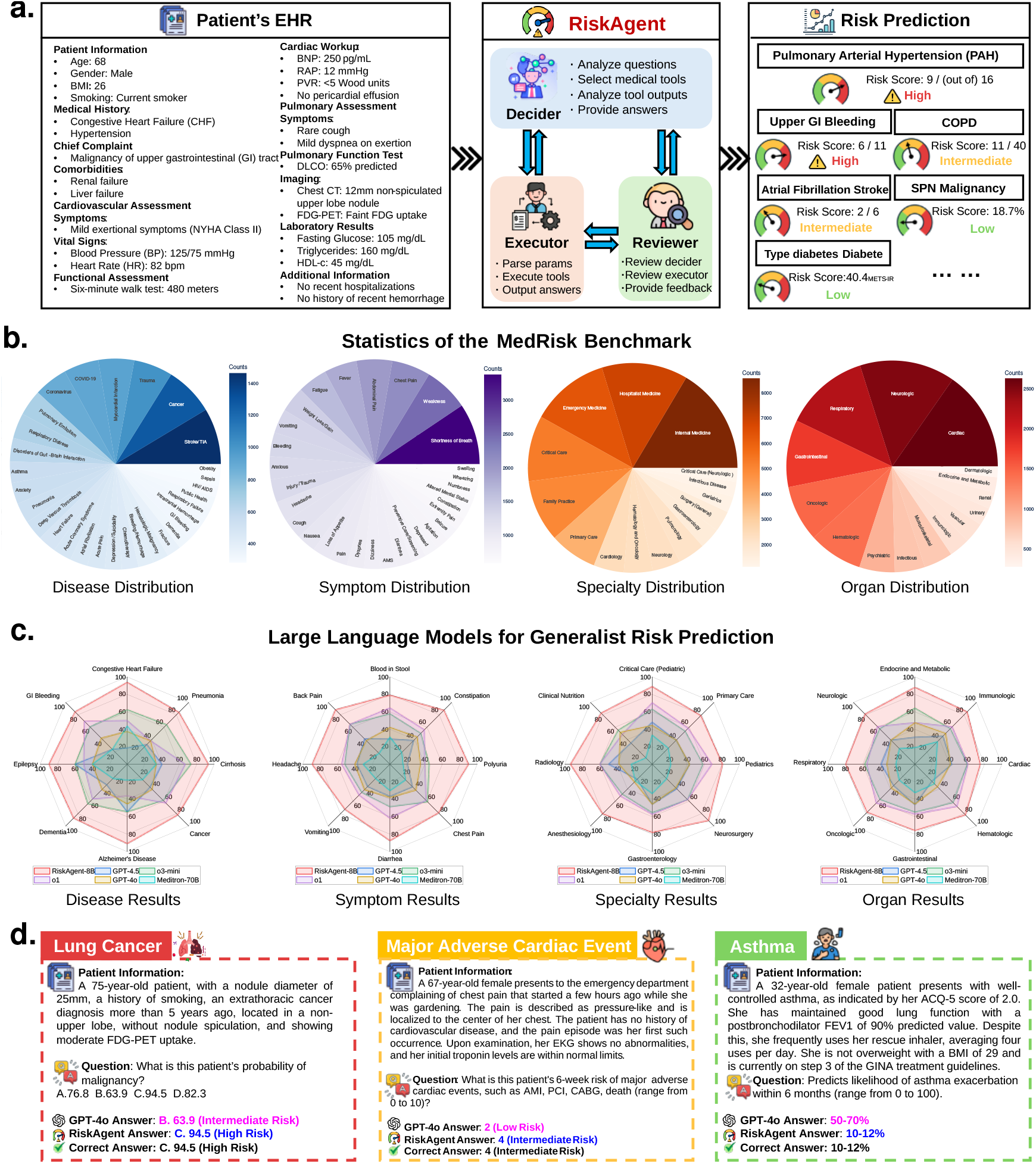
**a**. The RiskAgent, including three LLM agents (Decider, Executor, and Reviewer) can perform multiple medical risk predictions given the patient’s healthcare information. **b**. Statistics of the MedRisk benchmark, which consists of 12,352 medical risk questions, covering 154 diseases, 86 symptoms, 50 specialties, and 24 organ systems. **c**. With only 8 billion parameters, our RiskAgent outperforms both existing high-performance general LLMs (i.e., GPT-4o ^3^, o1^4^, o3-mini ^16^) and state-of-the-art medical LLM (i.e., Meditron-70B) by large margins across different diseases, symptoms, specialties, and organ systems. In contrast to existing LLMs, RiskAgent can collaborate with evidence-based medical tools to not only substantially increase its risk prediction accuracy, but also deliver evidence-based answers. The t-tests between the results from RiskAgent and the best-performing LLMs indicate that the improvement is significant with *p <* 0.01. **d**. The examples of risk predictions by our method for cancer, cardiac events, and asthma, demonstrate greater accuracy than GPT-4o. The pinkand blue-colored text indicates the incorrect and desirable answers, respectively.

Overall, our RiskAgent can make a wide range of risk predictions for hundreds of scenarios across diverse diseases. Our RiskAgent has the following advantages: 1) **Clinical efficacy**: The objective of an LLM is to learn to collaborate with existing medical tools supported by evidence-based medicine ^32^, rather than learning to perform various complex medical tasks. This greatly reduces LLM’s learning pressure and makes it easier to deliver superior performance and, therefore, is more efficient and straightforward than learning from a large volume of target task data as in conventional methods. **2. Resource efficiency**: In real-world settings, particularly hospitals, there are existing deployed medical tools, including devices, models, and APIs. Our method learns to collaborate with and directly utilize these tools, instead of designing and training new medical LLMs for clinical tasks, thus reducing resource costs and making our LLMs accessible for resource-limited clinical applications. This is a critical step toward sustainable AI. 3) **Privacy friendly**: The encouraging results of our model, with only 8 billion parameters, show the potential of our solution to build open-source LLMs in the real world with fewer privacy concerns compared to commercial LLMs. 4) **Faithful outputs**: Compared to existing methods, the collaboration with evidence-based medical tools not only brings more accurate performance, but also, most importantly, helps our model produce trustworthy results with means for clinicians to efficiently inspect the evidence of the output recommendations, which is critical in achieving trust from clinical users.

### Framework

Figure 2 shows the flowchart of our RiskAgent system. Inspired by the success of actor-critic reinforcement learning ^33^ that introduces the Critic, Actor, Reward, and Environment, we introduce three LLM agents and one component, i.e., Decider, Executor, Reviewer, and Environment. These modules work together to ensure that our system runs smoothly and accurately.

**Figure 2.**
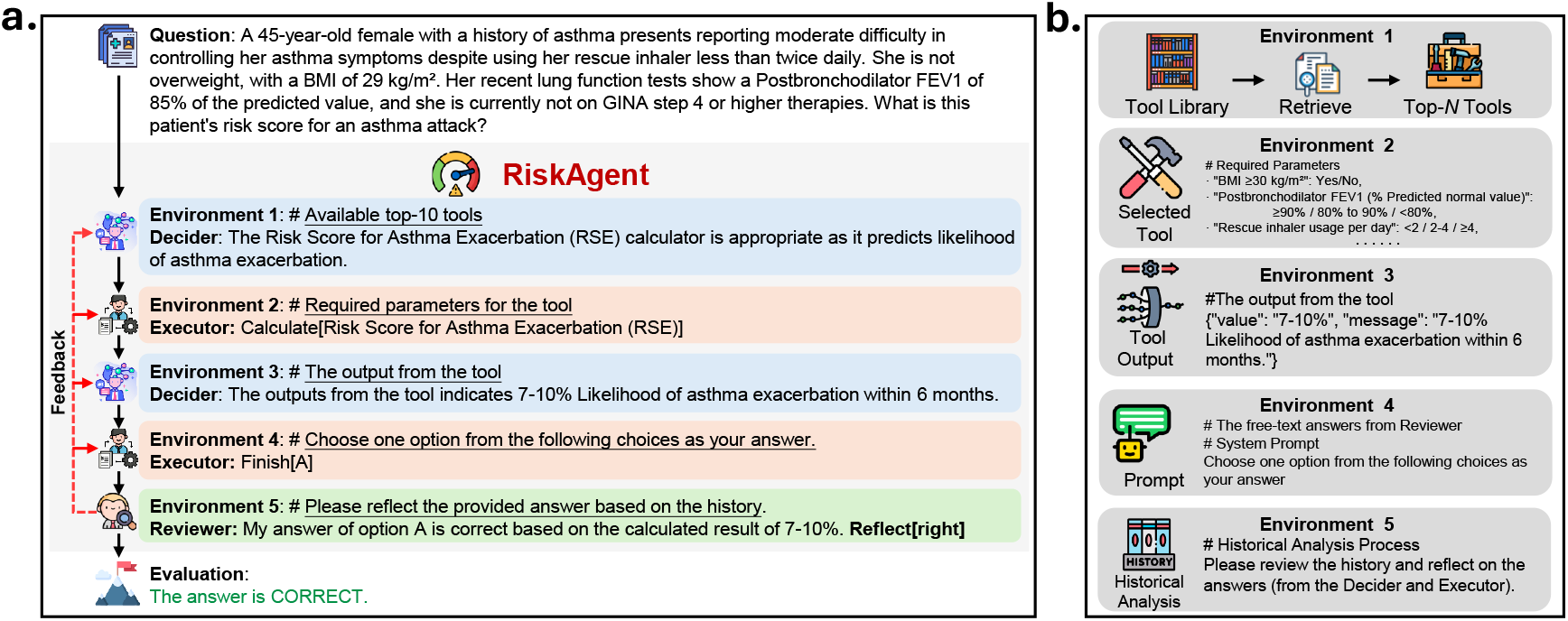
Flowchart of the RiskAgent system. **a**. Data flow in the system. **b**. Demonstration of the Environment component in the system.

- **Decider**: The LLM is instructed to analyze medical problems, decide which medical tools to invoke, analyze the output of the tools, and provide answers.
- **Executor**: The LLM is instructed to execute the Decider’s decisions, i.e., parse the required parameters to invoke the tools, and convert the answers into risk scores or choices for evaluation.
- **Reviewer**: The LLM is instructed to review all historical information and the provided answers, and then offer reflection results.
- **Environment**: It provides the system with external information that can be interacted with, such as the medical tool library, required parameters for the tools, tool outputs, and system prompts.

#### Decider-Executor

The Decider and Executor in the system are responsible for assigning and executing instructions, respectively. Their collaborative process is as follows: 1) The Decider, as shown in Figure 2, analyzes the given medical problem and the retrieved tools to determine which tools to use; 2) The Executor executes the Decider’s decisions (i.e., the determined tools). In detail, the Executor analyzes the medical problem according to the tools’ parameter requirements, parses the necessary parameters accurately, sends them to the tools for invocation, and obtains the tools’ results; 3) The Decider further analyzes the tools’ results reported by the Executor and generates answers to the input medical problems; 4) The Executor finally converts the generated answers into risk scores or risk groups as the system outputs. As we can see, our system reduces the learning burden of each component by dividing responsibilities between the Decider and Executor, thus enabling accurate and efficient collaboration with medical tools.

#### Reviewer-Decider/Executor

We further introduce the Reviewer that evaluates the provided answer by considering the historical analysis process and providing reflection results to improve the system’s performance. If the Reviewer determines that the answer is correct, i.e., Reflect[right], the system outputs the answer; otherwise, i.e., Reflect[wrong], the Reviewers provides reflection results to the Reviewer and Executor, and the system continues running. In the case of the Decider selecting the wrong tools, the Reviewer provides reflection results on the Decider’s decisions, and the Decider re-selects the tools. The Reviewer also handles the situation when the Executor fails to correctly implement the Decider’s decisions, such as a parameter parsing failure. The Reviewer, therefore, also reviews the progress of the Executor’s work to ensure the tool invocation is completed successfully.

### Environment

We introduce five environments to enhance collaboration between our system and medical tools, shown in Figure 2 (b).

- **Environment 1** Hundreds of medical tools are often involved in real-world clinical scenarios, due to their complexity and diversity. To enable LLMs to collaborate with these tools, we introduce a retrieval-ranking algorithm to extract the *N* most relevant medical tools from a tool library containing *M* tools based on the current medical problem.
- **Environment 2** When the Decider has chosen a tool, this environment provides the Executor with the required parameters for tool invocation, enabling the Executor to use the tools smoothly and accurately.
- **Environment 3** The returned results from the medical tools are stored in this environment, where the Decider interacts with for analysis and the generation of freetext answers to questions.
- **Environment 4** This environment provides the system prompts to assist the Executor in formatting and converting the free-text answers generated by the Decider into risk scores or choices for evaluation.
- **Environment 5** This environment stores the historical analysis process from the Decider and Executor, as well as system prompts that instruct the Reviewer to perform the review process accurately.

We attach a detailed description of our method in our supplementary material.

## Results

In this section, we comprehensively evaluate the effectiveness of our solution and compare it with current representative LLMs, covering both state-of-the-art closed-source commercial LLMs and open-source public LLMs with model parameters ranging from 7 to 70 billion.

### MedRisk Benchmark

We first build a benchmark MedRisk for evaluating LLM’s performance for risk prediction. To build an accurate benchmark dataset consisting of diverse risk problems, we first collect all available evidence-based tools, i.e., clinical calculators, from clinical-standard source MDCalc ^41^. We then review and exclude tools that are not directly predicting disease risks (e.g., those predicting Pregnancy Due Dates and BMI), retaining 387 tools that predict 154 diseases across different scenarios. We then use the APIs provided by MDCalc ^41^ to generate over 15,000 questions with free-text reference answers by requesting GPT-4o ^2^ in Azure to randomly select appropriate tool parameters, approximately 20 quantitative and 20 qualitative questions for each tool. Next, we manually identify and exclude data samples that are unrelated to risk prediction, obtaining 12,352 risk questions across 154 diseases, 86 symptoms, 50 specialties, and 24 organ systems. For robust and efficient evaluation, we convert these questions and free-text references to multiple-choice format by combining the correct answer with three wrong choices. Finally, we divide the benchmark into MedRisk-Qualitative and MedRisk-Quantitative, with 6,176 data samples each, according to their question and answer types. For example, the question ‘What is the estimated risk of postoperative pulmonary complications?’ (Answer: High risk) belongs to Qualitative, while the question ‘What is the Risk Score for Asthma Exacerbation (RSE) for this patient?’ (Answer: 30-37%) belongs to Quantitative. Figure 1(b) shows the statistics of the top-30 diseases, top-30 symptoms, top-15 specialties, and top-15 organ systems in the MedRisk benchmark.

To comprehensively evaluate LLMs’ performance for risk prediction, we collect 13 different types of representative LLMs and agents. 1) *Public LLMs*: We collect multiple representative open-source public LLMs across different numbers of model parameters, i.e., Mistral-7B ^34^, LLaMA-3-8B ^35^, Mixtral-8x7B ^36^, and LLaMA-3-70B ^35^; We also collect two public medical LLMs: PMC-LLaMA-13B ^11^ and Meditron70B ^15^. They are developed by fine-tuning general LLMs on a large corpus of medical datasets ^6^ to learn rich medical knowledge, achieving improved performance in medical tasks. 2) *Commercial LLMs*: We further collect the currently most powerful commercial LLMs, GPT-4o-2024-08-06^3^, o3-mini2025-01-31^16^, o1-2024-12-17^4^, and GPT-4.5^37^. 3) *AI Agents*: Recently, AI agents ^43,44^, which assign different LLMs to different roles to collaboratively perform tasks, have demonstrated better task performance than single LLMs in traditional domains such as gaming and programming. Therefore, for a comprehensive comparison, we further collect three wellknown and recent AI agents, i.e., ReAct ^38^, BOLAA ^39^, and Chameleon ^40^. We adopt LLaMA-3-8B ^35^ as the backbone of our system to re-implement them for a fair comparison.

### Main Evaluation

In Table 1, we report different methods’ performance on the most common cases across disease, symptom, specialty, and organ system and their overall performance. RiskAgent not only outperforms different types of LLM in all cases, respectively, but also achieves the highest overall accuracy of 78.34% and 76.33% in MedRisk-Qualitative and MedRisk-Quantitative, respectively. It indicates the robustness of our method across different diseases, symptoms, specialties, organ system, and question types, providing a more comprehensive risk prediction solution than previous methods. In detail, compared with large commercial LLMs (e.g., GPT-4o, GPT-o1, and GPT-o3-mini) and open-source LLMs (e.g., Meditron70B and LLaMA-3-70B), our RiskAgent outperforms them by *>*20.0% accuracy, with much (at least 10 times) fewer parameters. This is desirable in resource-limited medical settings. Our method also achieves the best results among AI agents, with accuracy surpassing previous methods by up to 35.83% overall accuracy.

To clearly understand the effectiveness of our approach, we visualize the results on MedRisk-Quantitative in Figure 1(c), which compares our method with the competitive LLMs. It shows that our RiskAgent outperforms previous methods in all cases. These encouraging results demonstrate that our RiskAgent achieves more precise and robust decision support than existing advanced LLMs for complex medical tasks. Importantly, our method uses evidence-based tools for prediction and traces the information source (i.e., evidence) behind our system’s decisions, making the results more transparent and reliable than existing approaches. In Figure 3, we further plot the results of five runs for RiskAgent-8B, GPT-4o, and o3-mini. We can see that RiskAgent-8B not only consistently achieves the best results across all cases in different runs but also achieves lower perturbations (STD) than o3-mini in most cases, especially in risk prediction for cardiac.

**Figure 3.**
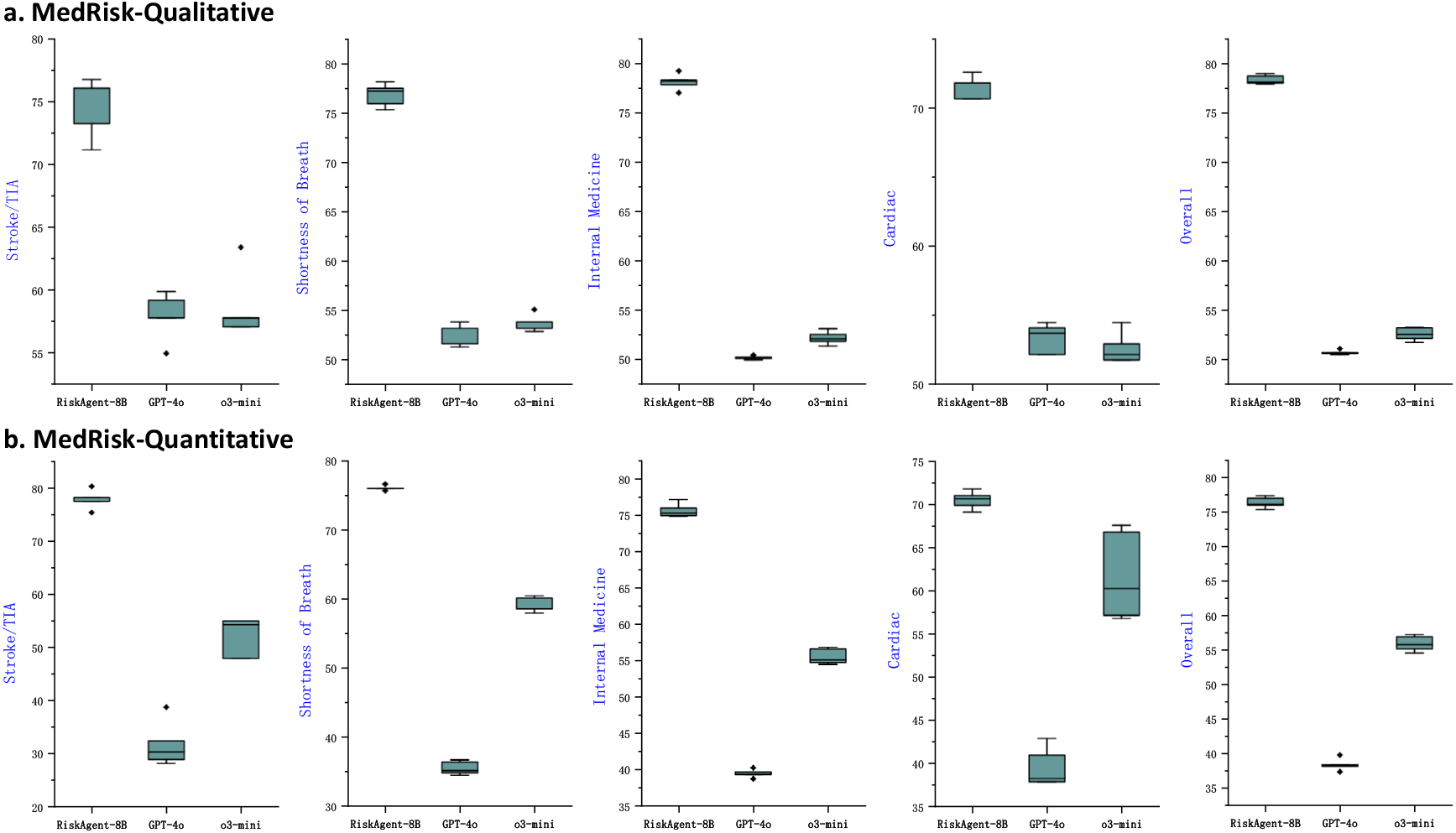
Performance for RiskAgent-8B, GPT-4o, and o3-mini. In the boxplot, the central line indicates the median value, while the lower and upper boundaries represent the 25th (Q1) and 75th (Q3) percentiles, respectively. The whiskers extend up to 1.5 times the interquartile range (IQR).

### Evaluation of Rare Diseases and Cancer

In this section, to evaluate our method’s robustness in solving diverse complex diseases, we further conduct experiments on rare diseases and cancer.

For rare (but important) diseases, we follow two clinical-standard rare disease databases, i.e., European Orphanet^1^ and the US National Organization for Rare Disorders (NORD)^2^, to choose five rare diseases for evaluation. The left part of Figure 4 illustrates the results of our RiskAgent and previous strong LLMs. As we can see, RiskAgent achieves the best results in all cases. In particular, in Amyloidosis, RiskAgent achieves greater improvements than the most recent commercial LLM, GPT-4.5. For cancer, as shown in the right part of Figure 4, we evaluate different methods on six cancers. We can clearly see that our method consistently achieves competitive results with different For example, on Gastric Cancer and Breast Cancer, RiskAgent surpasses previous LLMs by at least 37.5% and 28.57% accuracy, respectively.

**Figure 4.**
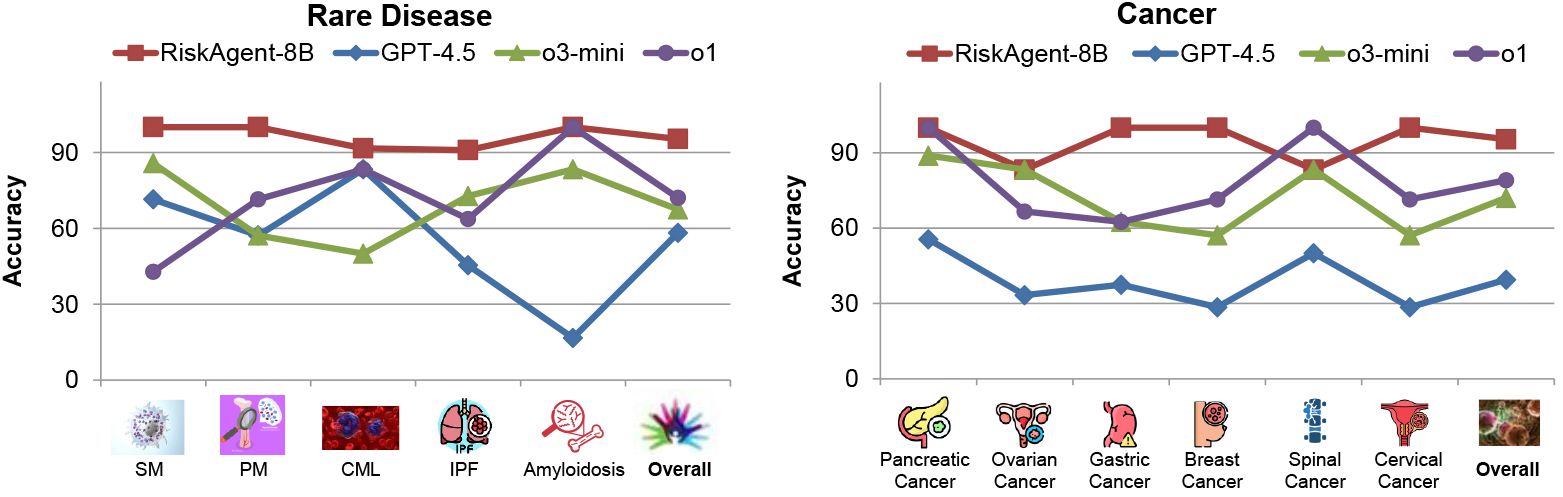
The robustness of our method: We evaluate the performance of models on five rare diseases (left) and six types of cancer (right). SM: Systemic Mastocytosis; PM: Primary Myelofibrosis; CML: Chronic Myelogenous Leukemia; IPF: Idiopathic Pulmonary Fibrosis.

Overall, our RiskAgent, with only 8B model parameters, achieves encouraging results across diverse rare diseases and cancers. It demonstrates the robustness of our method and shows the potential of our solution to provide more accurate and robust risk prediction for complex diseases, especially for rare diseases, compared to existing state-of-the-art LLMs.

### Generalization Analysis

In this work, we provide a solution that allows LLMs to collaborate with evidence-based medical tools to produce accurate, reliable medical results. Thus, our method is agnostic to the underlying LLM, making it adaptable to various LLMs and helping them produce accurate and evidence-based medical outputs. To this end, we further explore our method’s generalizability and applicability as LLMs continue to evolve rapidly over time and as the number of model parameters increases, especially considering the rapid development of LLMs and the trend toward increasing model sizes. During evaluation, we select the advanced commercial GPT-4o-series developed at different times (*gpt-4o-2024-05-13, gpt-4o-2024-08-06*, and *gpt-4o-2024-11-20*) and the representative public LLaMA-series (*LLaMA-3-1B, LLaMA-3-3B*, and *LLaMA-3-8B*), which share the model architecture but differ in model size.

We compare the performance of (i) the basic LLM and (ii) the basic LLM enhanced using our method. Figure 5 shows that our method provides significant performance improvements for all basic LLMs (accuracy of 17.04%*~*44.27%) on both MedRisk-Qualitative and MedRisk-Quantitative benchmarks. In detail, results on the GPT-4o-series indicate that as models evolve, the performance improvement brought by our method increases. This indicates that if downstream applications do not involve strict privacy regulations, our method can significantly improve the performance of large advanced LLMs, enabling them to accurately solve complex medical problems. Even when privacy concerns are involved, our method can still significantly boost the results of LLaMA-3-series public LLMs across different model parameters. For example, our method improves accuracy by 19.57% *~* 30.52% for the 1B and 3B LLMs, enabling them, with only 1B and 3B model parameters, to outperform the commercial GPT-4o basic LLMs. Such encouraging improvements in lightweight LLMs are useful in resource-limited medical institutions.

**Figure 5.**
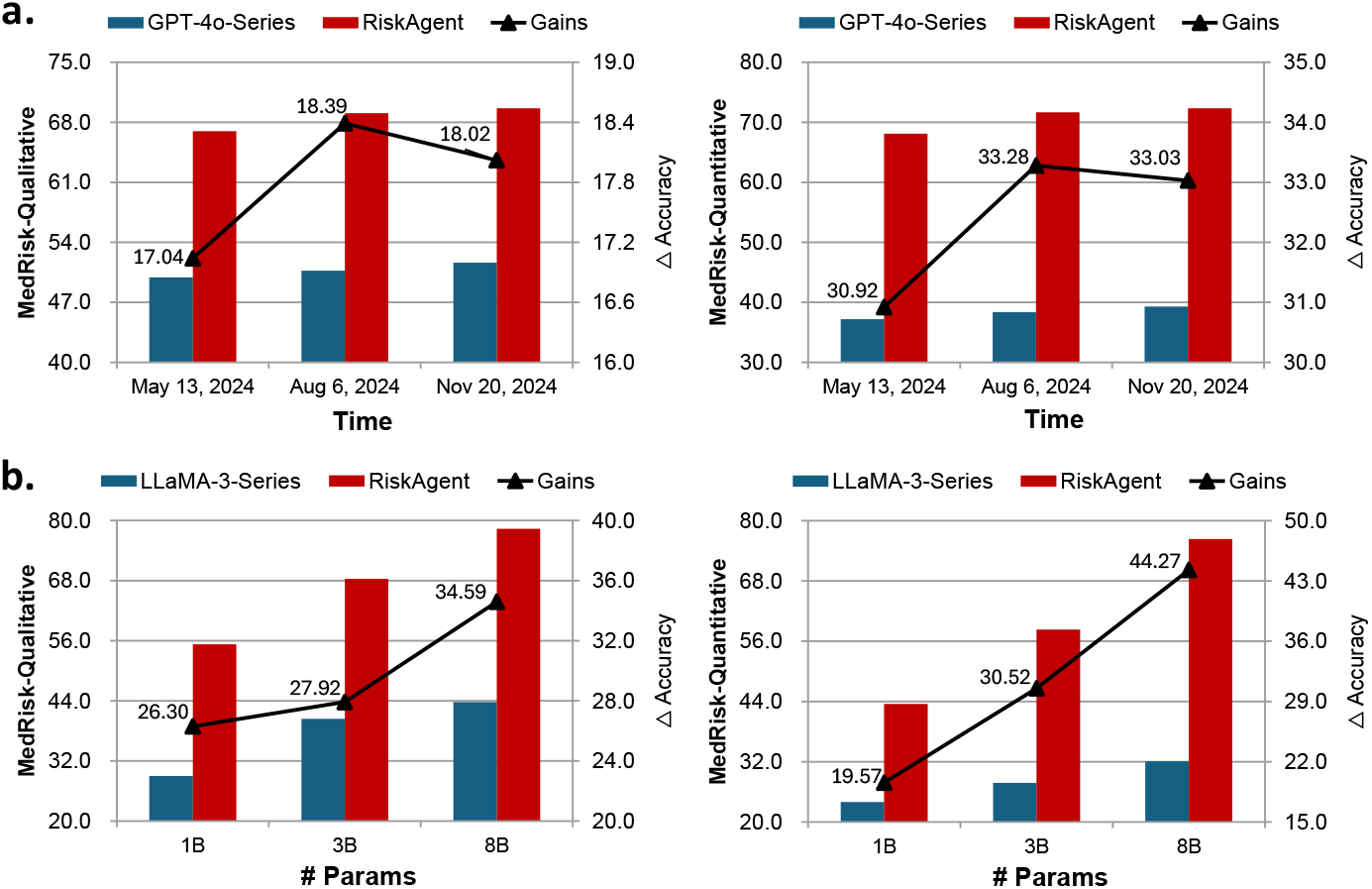
The generalization ability of our method: We report the overall accuracy of the basic LLMs (blue bars) and the basic LLMs enhanced using our method (red bars). We evaluate **a**. different variants of the GPT-4o-series LLMs developed at different times and **b**. LLaMA-3-series LLMs with varying numbers of model parameters. The polyline and the right y-axis show the improvements in different variants. We can see that the more advanced (**a**.) and the larger (**b**.) the basic LLM, the greater the improvements.

Meanwhile, we notice that as model sizes increase, the improvements brought by our method become greater. Overall, the improved results across different GPT-4o-series and LLaMA-3-series LLMs show the strong potential of our approach, enabling large and advanced LLMs to deliver accurate medical data analysis and evidence-based predictions. As LLMs continue to evolve rapidly, our method can further enhance their effectiveness in complex, critical medical tasks.

### External Evaluation

We further evaluate our method’s generalization ability on an external evidence-based diagnosis benchmark (MEDCALC-BENCH ^46^), which requires the models to use medical calculations specifically for clinical diagnosis to perform evidencebased decision support. Note that our RiskAgent is designed specifically for risk prediction; thus, this benchmark evaluates our methods’ generalization ability to new tasks and domains. For evaluation, we follow previous work ^46^ to employ zeroshot chain-of-thought (CoT) prompting to report the results. We adopt different backbones, i.e., LLaMA-3-8B ^35^ and GPT-4o ^3^, to implement two variants of our method: RiskAgent-8B and RiskAgent-GPT-4o. Table 2 demonstrates the outstanding performance of our approach on external evaluation using the evidence-based clinical benchmark MEDCALC-BENCH. The results clearly show that: (i) The RiskAgent-8B model, with only 8B parameters, surpasses all public LLMs, including the larger LLMs Meditron-70B ^15^ and Mixtral-8x7B ^36^ in major cases. (ii) The RiskAgent-GPT-4o model significantly outperforms all baseline models with an overall accuracy of 67.71%, representing a substantial improvement over the standard GPT-4 and GPT-4o. Most notably, our model exhibits exceptional performance in rule-based diagnosis tasks, achieving 61.67% accuracy in the Diagnosis category - more than double the performance of all baseline methods. Meanwhile, the performance improvements are particularly striking in calculation-heavy categories, with RiskAgent-GPT-4o achieving 97.08% on Physical assessments and 95.00% on Dosage calculations - areas where precise quantitative reasoning is critical for patient safety. It can also be verified by our RiskAgent-8B model, which, surprisingly, outperforms all public and commercial LLMs in dosage calculations. These encouraging results demonstrate that our RiskAgent framework not only excels in risk prediction, but also shows comfortable generalization capabilities across diverse diagnostic tasks in previously unseen clinical scenarios. The significant performance gains across both equation-based and rule-based calculations further underscore the robustness of our approach and its potential to provide reliable decision support across various medical contexts.

**Table 2.**
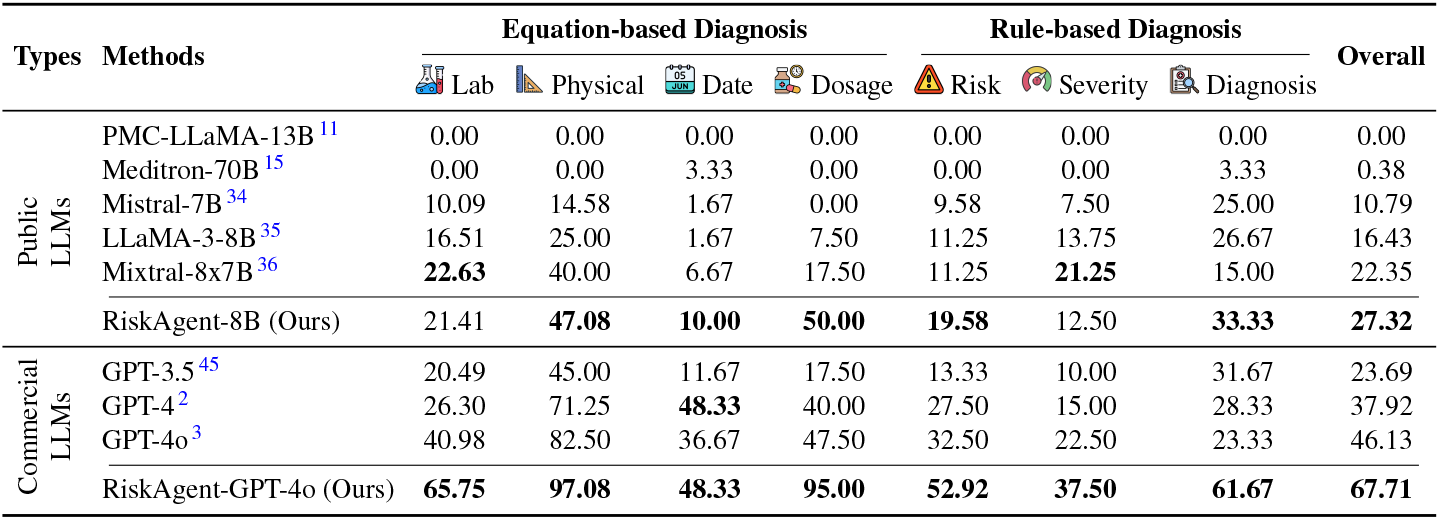
Evaluation of our RiskAgent, designed for risk prediction, on an external evidence-based clinical diagnosis benchmark (MEDCALC-BENCH ^46^), which is categorized into Equation-based (Lab, Physical, Date, Dosage) and Rule-based (Risk, Severity, Diagnosis) diagnosis. All values are reported in percentage (%).

| Types | Methods | Equation-based Diagnosis |  |  |  | Rule-based Diagnosis |  |  | Overall |
| --- | --- | --- | --- | --- | --- | --- | --- | --- | --- |
|  |  | Lab | Physical | Date | Dosage | Risk | Severity | Diagnosis |  |
| Public LLMs | PMC-LLaMA-13B<sup>11</sup> | 0.00 | 0.00 | 0.00 | 0.00 | 0.00 | 0.00 | 0.00 | 0.00 |
|  | Meditron-70B<sup>15</sup> | 0.00 | 0.00 | 3.33 | 0.00 | 0.00 | 0.00 | 3.33 | 0.38 |
|  | Mistral-7B<sup>34</sup> | 10.09 | 14.58 | 1.67 | 0.00 | 9.58 | 7.50 | 25.00 | 10.79 |
|  | LLaMA-3-8B<sup>35</sup> | 16.51 | 25.00 | 1.67 | 7.50 | 11.25 | 13.75 | 26.67 | 16.43 |
|  | Mixtral-8x7B<sup>36</sup> | **22.63** | 40.00 | 6.67 | 17.50 | 11.25 | **21.25** | 15.00 | 22.35 |
|  | RiskAgent-8B (Ours) | 21.41 | **47.08** | **10.00** | **50.00** | **19.58** | 12.50 | **33.33** | **27.32** |
| Commercial LLMs | GPT-3.5<sup>45</sup> | 20.49 | 45.00 | 11.67 | 17.50 | 13.33 | 10.00 | 31.67 | 23.69 |
|  | GPT-4<sup>2</sup> | 26.30 | 71.25 | **48.33** | 40.00 | 27.50 | 15.00 | 28.33 | 37.92 |
|  | GPT-4o<sup>3</sup> | 40.98 | 82.50 | 36.67 | 47.50 | 32.50 | 22.50 | 23.33 | 46.13 |
|  | RiskAgent-GPT-4o (Ours) | **65.75** | **97.08** | **48.33** | **95.00** | **52.92** | **37.50** | **61.67** | **67.71** |

### Ablation Study

In this section, we provide an ablation study in Table 3 to better understand the contributions of each component. Our method introduces four main components, i.e., the Tool Library in the environment, the Decider, the Executor, and the Reviewer, to achieve superior performance.

**Table 3.** Ablation study of our RiskAgent, which includes four main components: Tool Library, Decider, Executor, and Reviewer. We take LLaMA-3-8B ^35^ as the basic LLM and use accuracy to report our results. We add the results of a public LLM (LLaMA-3-70B ^35^) and a commercial LLM (GPT-4o ^3^) for comparison.

| Settings | Tool Library | Decider | Executor | Reviewer | MedRisk Qualitative |  |  | MedRisk Quantitative |  |  |
| --- | --- | --- | --- | --- | --- | --- | --- | --- | --- | --- |
|  |  |  |  |  | Tool Selection | Parameter Parsing | Overall Performance | Tool Selection | Parameter Parsing | Overall Performance |
| LLaMA-3-70B<sup>35</sup> | - | - | - | - | - | - | 48.62 | - | - | 34.42 |
| GPT-4o<sup>3</sup> | - | - | - | - | - | - | 50.68 | - | - | 38.39 |
| Basic LLM | - | - | - | - | - | - | 43.75 | - | - | 32.06 |
| (a) | ✓ | - | - | - | 82.06 | 35.06 | 57.95 | 78.49 | 32.71 | 46.59 |
| (b) | ✓ | ✓ | ✓ | - | 84.50 | 62.17 | 72.00 | 88.15 | 62.91 | 65.58 |
| RiskAgent-8B | ✓ | ✓ | ✓ | ✓ | **92.94** | **68.02** | **78.34** | **97.89** | **71.19** | **76.33** |

(i)The results show that all components contribute to improved performance, proving the effectiveness of each component. (ii) By comparing Basic LLM and Setting (a), which both MedRisk-Qualitative and MedRisk-Quantitative benchmarks. In detail, results on the GPT-4o-series indicate that as models evolve, the performance improvement brought by our method increases. This indicates that if downstream applications do not involve strict privacy regulations, our method can significantly improve the performance of large advanced LLMs, enabling them to accurately solve complex medical problems. Even when privacy concerns are involved, our method can still significantly boost the results of LLaMA-3-series public LLMs across different model parameters. For example, our method improves accuracy by 19.57% 30.52% for the 1B and 3B LLMs, enabling them, with only 1B and 3B model parameters, to outperform the commercial GPT-4o basic LLMs. Such encouraging improvements in lightweight LLMs are useful in resource-limited medical institutions.

Meanwhile, we notice that as model sizes increase, the improvements brought by our method become greater. Overall, the improved results across different GPT-4o-series and LLaMA-3-series LLMs show the strong potential of our approach, enabling large and advanced LLMs to deliver accurate medical data analysis and evidence-based predictions. As LLMs continue to evolve rapidly, our method can further enhance their effectiveness in complex, critical medical tasks.

### External Evaluation

We further evaluate our method’s generalization ability on an external evidence-based diagnosis benchmark (MEDCALC-BENCH ^46^), which requires the models to use medical calculations specifically for clinical diagnosis to perform evidencebased decision support. Note that our RiskAgent is designed specifically for risk prediction; thus, this benchmark evaluates our methods’ generalization ability to new tasks and domains. For evaluation, we follow previous work ^46^ to employ zeroshot chain-of-thought (CoT) prompting to report the results. We adopt different backbones, i.e., LLaMA-3-8B ^35^ and GPT- 4o ^3^, to implement two variants of our method: RiskAgent-8B and RiskAgent-GPT-4o. Table 2 demonstrates the outstanding performance of our approach on external evaluation using the evidence-based clinical benchmark MEDCALC-BENCH. The results clearly show that: (i) The RiskAgent-8B model, with only 8B parameters, surpasses all public LLMs, including the larger LLMs Meditron-70B ^15^ and Mixtral-8x7B ^36^ in major cases. (ii) The RiskAgent-GPT-4o model significantly outperforms all baseline models with an overall accuracy of 67.71%, representing a substantial improvement over the standard GPT-4 and GPT-4o. Most notably, our model exhibits exceptional performance in rule-based diagnosis tasks, achieving 61.67% accuracy in the Diagnosis category - more than double the performance of all baseline methods. Mean-while, the performance improvements are particularly striking in calculation-heavy categories, with RiskAgent-GPT-4o achieving 97.08% on Physical assessments and 95.00% on Dosage calculations - areas where precise quantitative reasoning is critical for patient safety. It can also be verified by our RiskAgent-8B model, which, surprisingly, outperforms all public and commercial LLMs in dosage calculations. These encouraging results demonstrate that our RiskAgent framework not only excels in risk prediction, but also shows comfortable generalization capabilities across diverse diagnostic tasks in previously unseen clinical scenarios. The significant performance gains across both equation-based and rule-based calculations further underscore the robustness of our approach and its potential to provide reliable decision support across various medical contexts.

### Ablation Study

In this section, we provide an ablation study in Table 3 to better understand the contributions of each component. Our method introduces four main components, i.e., the Tool Library in the environment, the Decider, the Executor, and the Reviewer, to achieve superior performance.

(i) The results show that all components contribute to improved performance, proving the effectiveness of each component. (ii) By comparing Basic LLM and Setting (a), which introduces the tool library to enable the basic LLM to use the evidence-based tool for risk prediction, we observe that Setting (a) significantly boosts the performance of the basic LLM by more than 14% accuracy, surpassing larger models like LLaMA-3-70B and GPT-4o. This proves the motivation and effectiveness of leveraging evidence-based tools to achieve improved performance on complex medical tasks. (iii) Considering that collaborating with tools involves both tool selection and parameter parsing for tool execution, we further report their accuracy. Comparing Setting (a) and Setting (b), we observe that introducing LLM agents can significantly boost performance—improving tool selection accuracy by 9.66%, parameter parsing accuracy by 30.20%, and overall task performance by 18.99%. This proves the effectiveness of our method in separating the collaboration process into tool selection and parameter parsing through the designed Decider and Executor. By distributing responsibilities, our approach reduces the model’s learning burden, enabling accurate and efficient collaboration with medical tools. (iv) Finally, we notice that the Reviewer further improves overall performance, showing the effectiveness of reflecting on the analysis process and thereby correcting potential errors. Such a reflection process is very similar to the ‘Aha moment’ in the recently popular DeepSeek-R1 model ^47^, further proving the effectiveness of our method.

Overall, the ablation study shows that both collaboration with evidence-based medical tools and the designed agents play an important role in helping LLaMA-3-8B outperform the most recent competitive commercial LLMs (i.e., o1, o3-mini, and GPT-4.5) with only 8 billion parameters.

## Data Availability

All data produced are available online at

https://github.com/AI-in-Health/RiskAgent

## Method

We now introduce our method in detail. As shown in Figure 1 (a) and Figure 2, our RiskAgent introduces three LLM agents and one component, i.e., Environment, Decider, Executor, and Reviewer.

### Environment

**Environment 1** identifies the most suitable tool from the tool library for subsequent calculations. In our work, we collect 387 tools *T* = *{t*_1_, *t*_2_, …, *t*_*M*_*}*, where *M* = 387 to form our tool library. To optimize tool selection, we present an embedding-based retrieval-ranking algorithm, which helps the method accurately and efficiently select the most appropriate tools relevant to patients from the massive available tools, thereby increasing overall model performance.

*Retrieval*. We first adopt the text-embedding-ada-002 model ^48^ to extract the embeddings *E*_*T*_ of all tools *T* using their tool metadata including both tool names and descriptions, with the dimension of 1,536, defined as follows:

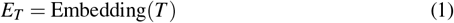

Then, we adopt the concatenation of patient information and risk questions to represent each patient. We again use the same embedding model to extract the embeddings *E*_*P*_ for all patients *P*.

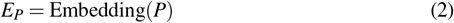

Finally, we adopt the widely-used cosine similarity to calculate the similarity scores *S* between patients and all available tools.

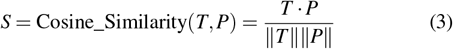

where *T · P* represents the dot product of the two vectors; and ∥*T*∥ and∥ *P*∥ are the Euclidean norms (i.e., *L*_2_ norms) of the vectors. Therefore, we can set a threshold and retrieve all relevant tools with a similarity score greater than it.

*Rank*. In our work, we propose ranking the tools using the calculated similarity scores *S* and forwarding the top-*N* tools with the highest similarity scores for the Decider to perform tool selection. We evaluate the performance of setting different values of *N*. Our preliminary results show that setting *N* = 5 achieves a 99.8% recall score, meaning that in 99.8% of cases, the correct tool is included among the top-5 tools retrieved and ranked by our method. This proves the effectiveness of the embedding-based retrieval-ranking algorithm used.

**Environment 2** provides structured parameter data so that an external medical tool can be invoked accurately. Once the tool is selected in Environment 1, this environment defines the required schema—including parameter names, types (numeric or categorical), and allowable value options—and ensures that each parameter in the patient information is extracted and validated in the correct format. Specifically, numeric fields must be cast appropriately, and categorical or boolean fields must match the tool’s defined options. This process guarantees that all required parameters are provided in a proper format, thereby minimizing the risk of miscalculation or incompatibility when the tool is invoked. To address the challenge of parsing inconsistencies in raw patient data, such as different unit systems, we provide a comprehensive input schema within our prompts. This schema explicitly specifies the required unit format (e.g., kilograms vs. pounds) and acceptable value ranges for each parameter. Additionally, we implemented a robust retry mechanism to handle potential parameter extraction or formatting errors. When inconsistencies are detected, the system prompts the LLM to re-analyze the patient data, guiding it to correctly extract and format the values according to the required specifications.

**Environment 3** manages how tool-generated results are interpreted and rephrased in light of the original patient information. After valid parameters are provided from Environment 2, the system invokes the chosen medical tool (e.g., DECAF Score) to calculate a risk level and yield a concise statement (e.g., “31% in-hospital mortality”). Environment 3 then transforms this raw output into a coherent free-text explanation that references both the patient information and the clinical question. For instance, a high-risk DECAF Score might be rephrased as: “A DECAF score of 4 places patients in a highrisk category, correlating with a mortality rate near 31%.” The Decider can then utilize this refined statement to guide higher-level decisions or generate a final answer.

**Environment 4** focuses on transforming high-level textual conclusions into standardized outputs, such as multiple-choice answers or discretized risk scores. When the Decider generates a preliminary conclusion in free-text form, this environment formats it into a concise, unambiguous response. For instance, in a multiple-choice setting, it explicitly requests the system to produce an answer in the form `Finish[A/B/C/D]`. This requirement helps ensure that the final output is straightforward to parse and evaluate, making it suitable for automated scoring against ground-truth labels.

**Environment 5** In the final stage of the pipeline, Environment 5 collects all historical analysis and system outputs, enabling the Reviewer to perform a global verification. The Reviewer checks the correctness of each step. If an error is detected, the Reviewer pinpoints the earliest failing environment and provides revision instructions. The system then re-executes from that stage, incorporating the feedback. This iterative cycle continues up to three attempts; if errors still persist beyond this limit, the process terminates with the last available result. Such a mechanism maintains overall computational efficiency while still allowing multiple opportunities to correct reasoning flaws.

### LLM Agents

#### Decider

The Decider is tasked with making key clinical decisions throughout the workflow. It first utilizes Environment 1 to identify and select the most contextually relevant tool, and then interacts with Environment 3 to interpret the outputs from that tool. Figure 6 (a) and (c) show the detailed prompts used in Reviewer. Leveraging the patient’s information and the tool’s computed results, the Decider produces a synthesized answer addressing the posed clinical question. This structured delegation—where the Decider focuses on reasoning and choice of tools—enhances clarity and reduces the complexity of subsequent tasks.

**Figure 6.**
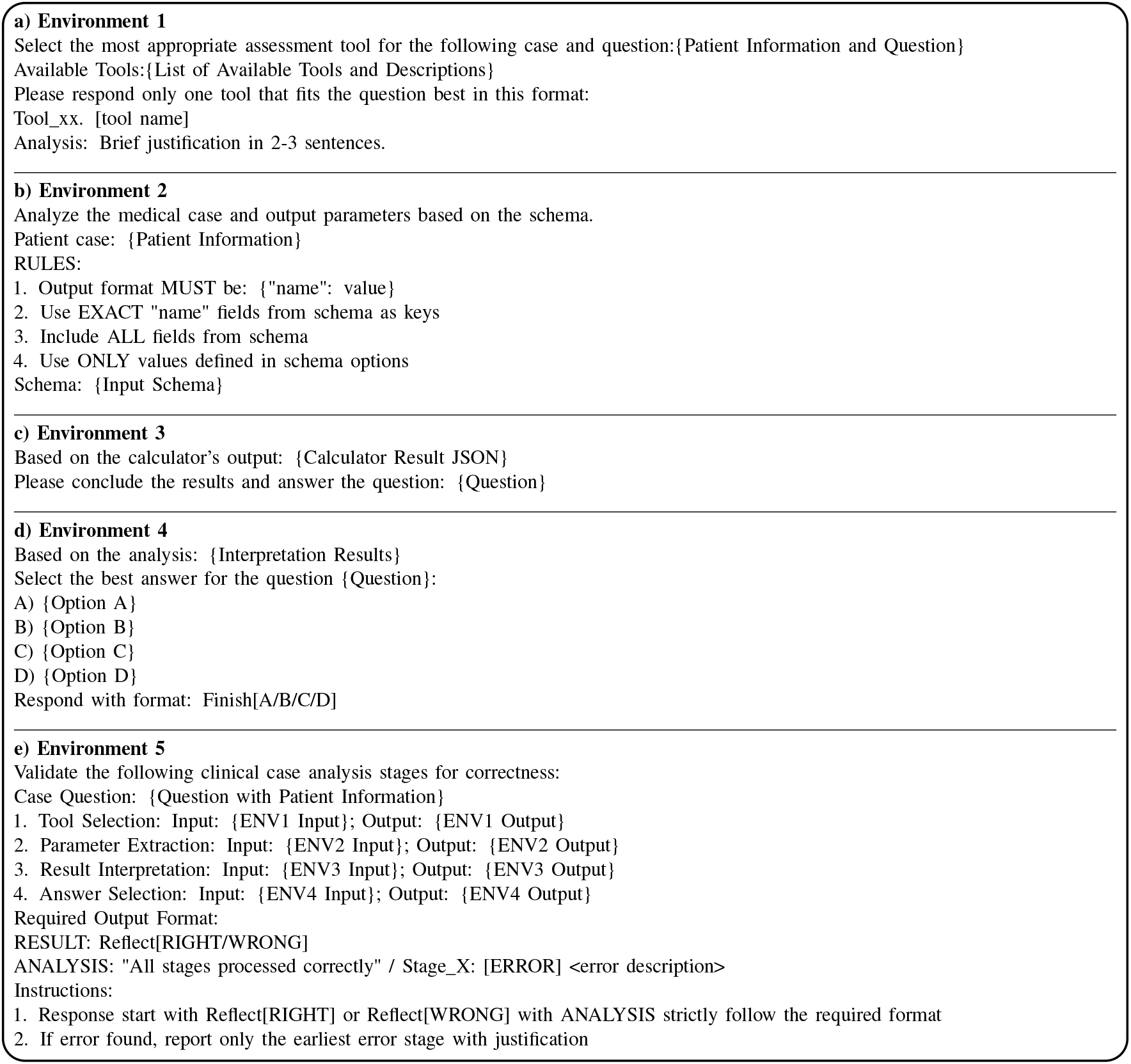
The instructions and prompts used in our three introduced LLM-based agents.

#### Executor

The Executor’s role is to carry out the instructions determined by the Decider precisely. After a specific tool is selected, the Executor references Environment 2 to extract and validate each input parameter. It then calls the tool externally and obtains its results. Figure 6 (b) illustrates the used system prompts. Finally, as shown in Figure 6 (d), the Executor consults Environment 4 to format the system’s textual output into a predetermined style (e.g., a discrete choice or numerical score).

#### Reviewer

The Reviewer, leveraging information stored in Environment 5, ensures the correctness and reliability of the entire pipeline. Once the Executor produces a final answer, the Reviewer meticulously inspects each environment’s input and output history. If any discrepancy or error is identified (e.g., using the wrong tool, misformatted parameters, or an incorrect conclusion), the Reviewer indicates the earliest environment that requires revision and provides targeted feedback. The system then restarts from that stage, integrating the Reviewer’s suggestions. Figure 6 (e) details the prompts used in Reviewer.

#### Instruction Fine-tuning

As we mentioned, enabling LLMs to accurately solve diverse medical tasks traditionally requires extensive medical data (ranging from 1 million to 80 billion tokens ^15,11,49,20,21,8^) and substantial computational resources that may be prohibitively expensive for fine-tuning.

Following the workflow established in Environments 1-5, we construct samples with explicit instructions and welldefined output formats. Each sample includes: (a) patient information, clinical queries, and candidate tools, with the expected output being the selection of the correct tool with justification; (b) patient information and tool schema, with the expected output being correctly formatted parameter extraction; (c) the original results computed by calculators with the sample question, with the expected output being a rephrased answer to the question; (d) outputs from step c along with questions and options, with the expected output being the final selected option; and (e) information from steps a-d, with the expected output being a comprehensive reflection (the instruction template is shown in Figure 6). We take the LLaMA-3-8B LLM ^35^ as our backbone to train our model using parameter-efficient fine-tuning techniques (LoRA) ^50^, significantly reducing computational requirements while maintaining high performance across medical risk prediction tasks.

It is worth noticing that we only train a single LLaMA-3-8B model for all the three agents across five environments rather than developing separate model for each. This ensures that the model parameters of our system is 8B making it fairly comparable to existing methods and LLMs.

### Experiment Settings

We perform model fine-tuning using LoRA ^50^ following the approach introduced using the LLaMA Factory framework ^51^. The MedRisk benchmark dataset is divided in a 7:1:2 ratio for training, validation, and testing, respectively. All training is conducted using PyTorch’s *DistributedDataParallel* on 4×A100 (80GB) GPUs with BF16 precision. We use AdamW optimizer ^52^ with a cosine learning rate scheduler. The effective batch size is 32 (batch size 8 per device with 4 gradient accumulation steps). The detailed hyper-parameters for training are shown in Table 4.

**Table 4.** We illustrate the detailed hyper-parameters for model training.

| Hyper_parameters | Value |
| --- | --- |
| GPUs | 4*A100 (80GB) |
| epochs | 5 |
| learning_rate | 0.0001 |
| model_max_length | 2,560 |
| per_device_batch_size | 8 |
| gradient_accumulation_steps | 4 |
| batch_size | 8*4*4 |
| warmup_ratio | 0.1 |
| optimizer | AdamW |
| lr_scheduler | cosine |
| precision | bf16 |
| finetuning_method | LoRA |
| lora_r | 8 |
| lora_alpha | 16 |
| lora_dropout | 0 |
| lora_target_modules | all |

During evaluation, for a fair comparison, we test all models on the same test set using a consistent protocol. We use a temperature of 0.6 and top-p of 0.9 for controlled randomness in generation. Each model receives identical input formatting, consisting of patient information, a clinical question, and four multiple-choice options. The evaluation metrics focus on accuracy, calculated as the exact match between model predictions and ground truth answers.

#### Ethical considerations

Our study was conducted on retrospective, de-identified data that fell outside the scope of institutional review board oversight. For the public data, all necessary patient/participant consent has been obtained, and the appropriate institutional forms have been officially archived. The clinical reader study component of this research involved the participation of licensed clinicians.

## Data Availability

The data used in our work is available at https://github.com/AI-in-Health/RiskAgent

## Code Availability

The code that supports the findings of this study is available at https://github.com/AI-in-Health/RiskAgent.

## Acknowledgements

This work was supported in part by the Pandemic Sciences Institute at the University of Oxford; the National Institute for Health Research (NIHR) Oxford Biomedical Research Centre (BRC); an NIHR Research Professorship; a Royal Academy of Engineering Research Chair; the Well-come Trust funded VITAL project; the UK Research and Innovation (UKRI); the Engineering and Physical Sciences Research Council (EPSRC); the InnoHK Hong Kong Centre for Cerebro-cardiovascular Engineering (COCHE); and the Clarendon Fund and the Magdalen Graduate Scholarship.

The Applied Digital Health (ADH) group at the Nuffield Department of Primary Care Health Sciences is supported by the National Institute for Health and Care Research (NIHR) Applied Research Collaboration Oxford and Thames Valley at Oxford Health NHS Foundation Trust. The views expressed are those of the author(s) and not necessarily those of the NHS, the NIHR or the Department of Health and Social Care.

## Author Contributions

F.L. conceived and designed the study. F.L. and J.W. contributed to the data analysis, implementations, and experiments, and prepared the manuscript. All authors contributed to the final manuscript preparation.

## Footnotes

1 https://www.orpha.net/

2 https://rarediseases.org/

